# Cumulative and relativistic temperature metrics for public heat alerts: A new approach proposed for greater Vancouver, Canada

**DOI:** 10.1101/2025.09.05.25333794

**Authors:** Alexi T. Hu, Michael Brauer, Eric Lavigne, Sarah Henderson

## Abstract

**Background:** Many heat alert systems rely on fixed absolute temperature thresholds that may not fully characterize risk. Furthermore, absolute temperatures can vary widely across some urban areas, leading to the same absolute thresholds being associated with different risks. The impending modernization of meteorological services in Canada provides an opportunity to address these challenges with heat alerts in greater Vancouver, British Columbia (BC).

**Objectives:** (1) To evaluate a cumulative heat alert indicator based on the sum of high temperatures over two consecutive days and the intervening overnight low (High + Low + High, or H+L+H) using mortality and heat-related morbidity data. (2) To compare the performance of absolute and relativistic H+L+H thresholds for generating heat alerts, including consideration of susceptible populations.

**Methods:** Time-series analyses were used to examine the relationships between all-cause mortality (2008-2024), heat-related emergency department (ED) visits (2014-2023), and H+L+H temperature observations from three weather stations across greater Vancouver. For each station, the outcomes were modelled using absolute and relativistic H+L+H thresholds, including analyses stratified by age, sex, socioeconomic status, and chronic health conditions.

**Results:** The relativistic H+L+H indicator was associated with significant risk of morbidity and mortality, with similar exposure-response functions for all three weather stations. In contrast, absolute thresholds showed considerable heterogeneity. The relativistic metric also showed different risk functions for some susceptible groups, including those with schizophrenia, Parkinson’s disease, and receiving income assistance.

**Conclusion:** The relativistic H+L+H temperature metric integrates cumulative multi-day exposure and addresses the challenge of intra-urban variability in absolute temperatures. This approach provides a more flexible alternative to heat alerts based on absolute temperatures, and highlights differences in risk among susceptible populations.

## Introduction

Climate change is increasing the frequency, intensity, and duration of extreme heat events (EHEs) globally^1–4^. In 2024, global mean surface temperatures reached approximately 1.55°C (±0.13°C) above pre-industrial levels (1850-1900)^5^. In British Columbia (BC), Canada, warming has outpaced global averages, where temperatures increased by up to 1.9°C between 1948 and 2016^6^. This regional warming has intensified the severity of EHEs, most dramatically illustrated by the 2021 event in western North America, during which the BC village of Lytton recorded a national temperature record of 49.6°C before its destruction by wildfire the following day^7,8^.

The growing severity of EHEs poses substantial public health threats and has led to major mortality and morbidity events worldwide. The European 2003 heatwave, Russia’s 2010 event, and the 2010 India-Pakistan heatwave demonstrate the deadly potential of sustained high temperatures^9–12^. In BC, the unprecedented 2021 EHE resulted in approximately 740 excess deaths^13^, and 619 deaths were directly attributed to the EHE by the BC Coroners Service^14^.

These events exacerbate cardiovascular disease, respiratory disease, mental illness, and other health conditions, overwhelm healthcare services, and disproportionately impact elderly, chronically ill, and socioeconomically disadvantaged populations^15–21^.

In response, many regions have established heat-health warning systems, that use predetermined temperature thresholds or meteorological conditions to trigger heat alerts and public health interventions^22–28^. In BC, the first regional Heat Alert and Response System (HARS) was established for greater Vancouver after the deadly 2009 EHE^29,30^. New temperature criteria were introduced in 2018, when Environment and Climate Change Canada (ECCC) first expanded heat alerts to cover the entire province^31^ and the provincial HARS was developed following the 2021 EHE^32^.

Like many heat warning systems globally, the BC HARS currently relies on fixed absolute temperature thresholds to issue heat alerts, requiring specific conditions (e.g., consecutive daytime maximum and overnight minimum temperatures) to be simultaneously met^23–28,33,34^. For example, the criteria for greater Vancouver are 29°C-16°C-29°C, meaning that two consecutive days must have forecasted high temperatures ≥29°C with an intervening overnight low ≥16°C for a heat alert to be issued. While conceptually and operationally straightforward, this approach may miss potentially hazardous conditions that do not meet the criteria.

Here we propose that the cumulative sum of two daily high (H) and one overnight low (L) temperatures may provide a more flexible framework for heat alerts. Using this approach, the current criteria for greater Vancouver would translate to a H+L+H value of 74°C. As an example, two scenarios that sum to 74°C are 28°C-18°C-28°C or 27°C-16°C-31°C, neither of which would trigger alerts under current fixed criteria. We evaluate the H+L+H approach using regional all-cause mortality and heat-related morbidity data, comparing results using absolute and relativistic values measured at three weather stations across greater Vancouver. Finally, we explore how heat alerts based on relativistic H+L+H criteria are associated with health outcomes across susceptible subpopulations.

## Methods

### Study area

The urban greater Vancouver area is located on the southwestern coast of BC (eFigure 1), with approximately 2.97 million residents representing nearly 60% of the provincial population according to the 2021 Canadian Census^35,36^. The region has a temperate oceanic climate characterized by mild, wet winters and warm, relatively dry summers^37^. Average summer daytime temperatures typically range from 17°C to 22°C^38^, but have been increasing in recent years^6^. Coastal areas generally experience cooler conditions than inland areas due to moderating ocean breezes^37^. Given historically mild summers, access to home air conditioning remains limited in greater Vancouver. According to Statistics Canada, only 32% of BC households had any type of air conditioning in 2023, with disproportionately lower access among racialized communities, renters, and low-income households^39^.

### Platform for Analytics and Data (PANDA)

This study used the Provincial Health Services Authority Platform for Analytics and Data (PANDA), which evolved from BC’s COVID-19 Data Library^40^, to link individual-level data across multiple administrative databases. The single-payer healthcare system in BC ensures comprehensive health data collection for individuals covered by the provincial plan. Multiple health services datasets were linked through a unique, anonymous patient master key. This study was conducted in the data mart titled “Evaluation of 2021 heat dome and extreme cold impacts in BC” (nb0107aa).

### Mortality and morbidity data

We extracted daily all-cause mortality records from the BC vital statistics database from May to September of 2008-2024. These data include the date of death, primary and secondary causes of death coded according to the International Classification of Diseases, 10^th^ Revision (ICD-10), age, sex, health region, and setting of death (e.g., acute care, long-term care, or in the community). Daily mortality counts were aggregated into rolling three-day sums (n_t-1_ + n_t_ + n) to smooth the data and maintain consistency with prior analyses^31,40^.

We extracted daily emergency department (ED) visit records from the National Ambulatory Care Reporting System (NACRS) for May to September of 2014-2024. Heat-related ED visits were identified using the five diagnostic and complaint codes used for provincial public health surveillance: general weakness, syncope/pre-syncope, altered level of consciousness, heat-related issues, and effects of heat and light. The ED visits were aggregated over three days in the same way as the mortality data.

### Comorbidities

Comorbidities among those who died or visited the ED were identified using the Chronic Disease Registries (CDRs), which track the incidence and prevalence of 26 chronic conditions using administrative case definitions^41^. For each study subject, we included binary flags to indicate the presence of heart failure, ischemic stroke, acute myocardial infarction, schizophrenia, substance use disorder, chronic obstructive pulmonary disease (COPD), parkinsonism, chronic kidney disease, and other chronic conditions. A multimorbidity measure, defined as the number of chronic conditions per individual, was also calculated.

The CDR is updated every fiscal year (April through March) in BC, and CDR data were only available until 30 March 2024 at the time of analysis. As such, deaths and ED visits in the summer of 2024 could only be linked to the CDR up to 30 March 2024 using the patient master key. To ensure all deaths and ED visits were treated equally in the sub-group analyses, the CDR up to 30 March of each year was used for all assessments. For example, deaths and ED visits in summer 2020 were matched the 2019-2020 fiscal year CDR.

### Income assistance

We used the PharmaNet database to identify patients enrolled in PharmaCare Plan C. PharmaNet tracks all medications dispensed by community and hospital outpatient pharmacies in BC^42^, and Plan C fully covers prescription costs for low-income residents receiving social benefits^43^.

Prescription records for one year preceding each death or ED visit were searched. Individuals with one or prescriptions covered by Plan C during this period were classified as low-income, those without any prescriptions were classified as unknown coverage, and all others were not covered by Plan C.

### Temperature

Daily temperature observations for May-September (2008-2024) were extracted from ECCC weather stations in Vancouver, White Rock, and Abbotsford, which are all municipalities of the greater Vancouver area. These stations are on the far west coast, slightly inland, and fully inland, respectively (eFigure1). Temperatures were recorded in integer degrees Celsius. We derived the cumulative 2-day temperate metric by summing the daily high measurements with the intervening overnight low (H+L+H). Relative percentiles of the H+L+H metric were calculated based on the 2008-2024 distribution of observed H+L+H values at each of the three stations. In all analyses, temperature data from the different weather stations were applied to the same health outcomes data for the whole region.

### Statistical analyses

All analyses were conducted using R version 4.5.0^44^. Negative binomial generalized additive models (GAMs) were first used to select 5 knots per calendar year to control for long-term time trends, based on minimization of the Akaike Information Criterion (AIC). Distributed lag non-linear models (DLNMs)^45^ were then used to identify the minimum mortality or morbidity values for the H+L+H metric at each weather station. We specified natural cubic splines with 4 degrees of freedom for temperature and a 3-day lag period using cross-basis functions in the DLNMs.

The minimum values were defined as the H+L+H associated with the lowest cumulative odds ratios (ORs) predicted by the DLNMs across the range of H+L+H absolute values for each outcome and weather station. Corresponding H+L+H percentile values were then derived from the absolute H+L+H values across the entire study period.

To quantify health risks above absolute and relative H+L+H thresholds, we used negative binomial GAMs to model relationships at values higher than the minimum mortality and morbidity thresholds, with data below the minimums serving as the reference category. For absolute H+L+H values, we modeled every integer degree above the minimum values. For relativistic values, we modeled each percentile above the minimum up to the 98th percentile, and then in 0.2-percentile increments up to the 99.8th percentile.

### Subgroup analyses

We fitted negative binomial models to assess the performance of the relativistic H+L+H approach across population subgroups by age, sex, place of death, PharmaCare Plan C coverage, and the presence of specific chronic conditions or multimorbidity. These subgroup analyses aimed to evaluate whether the relativistic method effectively captured varying heat-related risks among different demographic and clinical subpopulations.

### Case study of current versus proposed approach

Using data from the White Rock weather station, we identified days that met the current absolute BC HARS heat alert criteria (29°C-16°C-29°C) and those that met the equivalent H+L+H value (≥74°C). Dates meeting either criterion were compared visually and used to estimate risk of all-cause mortality and heat-related morbidity. Negative binomial GAMs were used to estimate ORs across three subsets of dates: those identified by the current criteria, those missed by the current criteria but captured by the H+L+H criteria, and all days ≥74°C.

### Sensitivity analyses

We conducted two sensitivity analyses. First, we applied alternative model specifications, including using 8 degrees of freedom for the temperature spline, a 7-day lag structure, and 3 knots per year to account for long-term trends. Second, we repeated all analyses using temperature forecasts issued three days in advance instead of temperature observations, given that heat alerts are always based on forecast data. Specifically, temperature forecasts for May through September (2008-2024) were obtained for Vancouver and Abbotsford, and the cumulative H+L+H values were calculated. Percentiles of the forecast H+L+H values were derived from the observed H+L+H distributions. Forecast data from White Rock were excluded because they were only available from 2019 onward.

## Results

### Population characteristics

This study included 122,050 all-cause deaths (2008-2024) and 196,754 heat-related ED visits (2014-2024) in greater Vancouver during May-September. Most deaths occurred among adults aged ≥65 years (79%), predominantly in hospitals or residential care settings (79%). In comparison, ED visits had a younger age distribution, with 56% among people aged ≥65 years.

Females comprised 47% of deaths and 50% of ED visits. Among decedents, 9% had at least one prescription dispensed under Plan C in the previous year, and this increased to 16% among ED visits. Most study subjects had at least one pre-existing chronic conditions and over 80% had three or more conditions. The most prevalent conditions included heart failure, chronic kidney disease, COPD, and substance use disorder (Table 1).

**Table 1.** Summary statistics for all-cause mortality and heat-related emergency department (ED) visits. Total sample sizes are presented for both main outcomes and comorbidity-specific outcomes. Proportions are reported for all subgroups.

|  | All-cause mortality | Heat-related ED visits |
| --- | --- | --- |
| **N** | 122,050 | 196,754 |
| **Age** |  |  |
| Under 65 | 21.2% | 43.4% |
| 65 or above | 78.8% | 56.5% |
| **Location of death** |  |  |
| In community | 21.3% | - |
| In care facility | 78.7% | - |
| **Sex** |  |  |
| Female | 47.5% | 50.3% |
| Male | 52.5% | 49.7% |
| **PharmaCare plan** |  |  |
| Plan C | 8.7% | 16.0% |
| Other plans | 83.0% | 77.6% |
| Unknown | 8.3% | 6.4% |
| **Comorbidities** |  |  |
| Any | 91.1% | 86.6% |
| Three or more | 81.5% | 84.3% |
| Heart failure | 24.0% | 22.4% |
| Ischemic stroke | 6.2% | 6.5% |
| Acute myocardial infarction | 9.2% | 8.8% |
| Schizophrenia | 3.5% | 5.6% |
| Substance use disorder | 13.2% | 19.6% |
| Chronic obstructive pulmonary disease | 19.1% | 16.5% |
| Parkinsonism | 2.9% | 3.3% |
| Chronic kidney disease | 22.5% | 29.3% |
| Other chronic conditions | 33.5% | 28.7% |

### Minimum mortality and morbidity H+L+H values

Between 2008 and 2024, the mean cumulative H+L+H values were 52.9°C in Vancouver, 53.0°C in White Rock, and 56.1°C in Abbotsford (eTable 1). DLNMs identified minimum mortality at 60°C for Vancouver and White Rock, and 70°C for Abbotsford. For ED visits, minimum values were at 55°C in Vancouver, 65°C in White Rock, and 70°C in Abbotsford (Figure 1). To promote consistency across models, we used H+L+H <60°C as the reference category for all analyses. This threshold corresponded to the 68th percentile in Vancouver, 66th percentile in White Rock, and 64th percentile in Abbotsford. For relativistic analyses, data below the 68th percentile served as the reference category.

**Figure 1.**
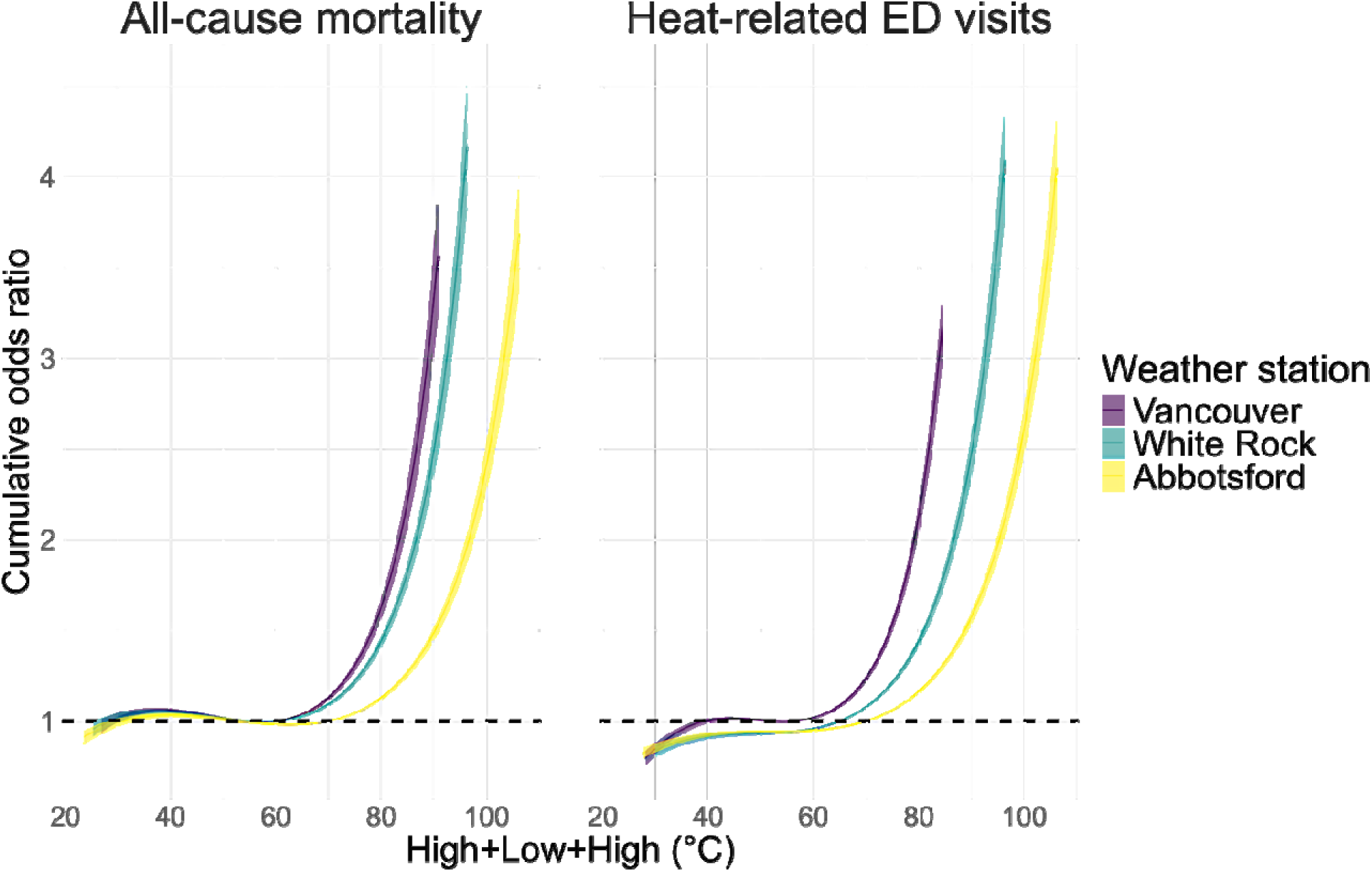
Exposure-response curves showing the associations between the High + Low + High (H+L+H) temperature indicator and all-cause mortality (left) and heat-related emergency department (ED) visits (right) using data from the three weather stations.

### Absolute versus relativistic H+L+H thresholds

Using absolute H+L+H values from the three weather stations, the exposure–response curves varied markedly. Risk of mortality increased sharply above 74°C using the Vancouver and White Rock temperatures, whereas the increase was more gradual for the Abbotsford data (Figure 2).

**Figure 2.**
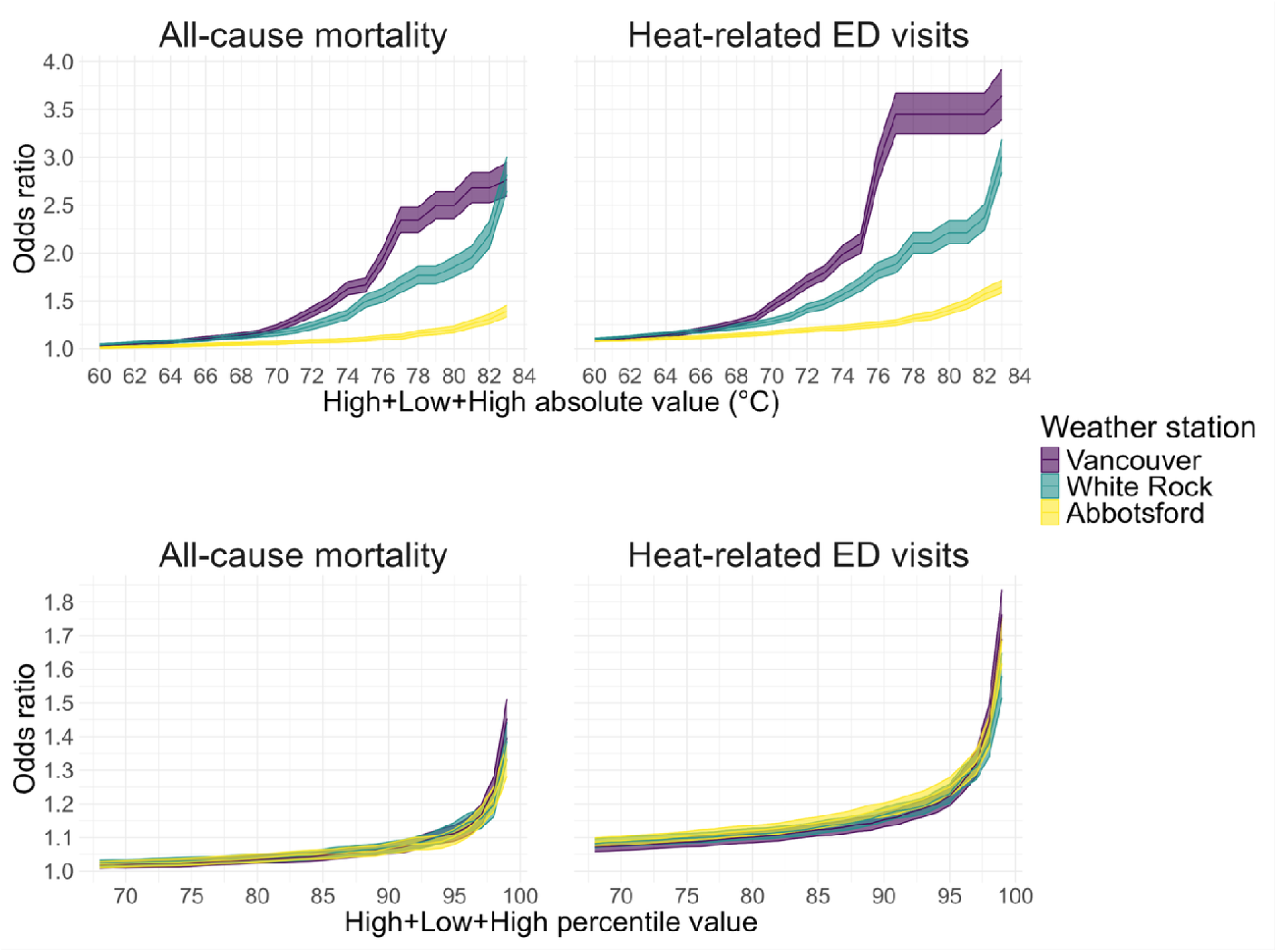
Exposure-response curves for the absolute (top) and relativistic (bottom) High + Low + High (H+L+H) temperature values and all-cause mortality (left) and heat-related emergency department (ED) visits (right) across three weather stations in greater Vancouver.

At H+L+H values ≥74°C the mortality ORs [95 % confidence interval] were 1.63 [1.56, 1.70] fo Vancouver, 1.35 [1.30, 1.40] for White Rock, and 1.09 [1.07, 1.11] for Abbotsford.

By contrast, the relativistic H+L+H values produced more consistent exposure–response relationships across all three stations, with similar shapes and inflection points. Risk increased gradually from the 68th to the 95th percentile, followed by steeper rises beyond the 95th percentile (Figure 2). At or above the 95th percentile, the mortality ORs were 1.11 [1.09, 1.14] for Vancouver, 1.12 [1.10, 1.15] for White Rock, and 1.11 [1.08, 1.12] for Abbotsford.

### Subgroup analyses

The relativistic H+L+H approach was applied to mortality and morbidity subgroups using temperature data from the White Rock weather station, comparing outcomes ≥95th percentile with those <68th percentile reference category. These analyses showed variations in heat-related susceptibility across subpopulations with chronic conditions (Figure 3) and other susceptibilities (Figure 4).

**Figure 3.**
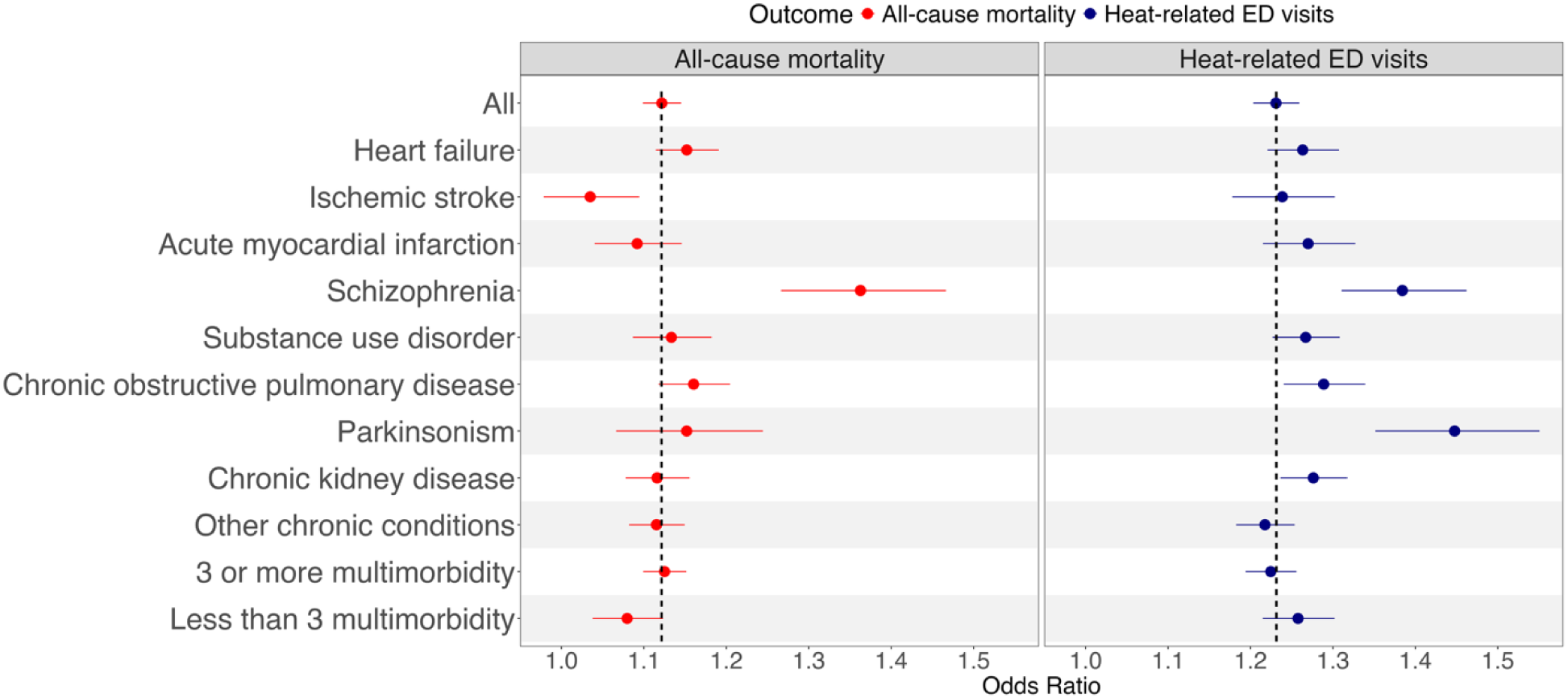
Odds ratios for all-cause mortality and heat-related emergency department (ED) visits, stratified by chronic conditions, using White Rock temperature data with High + Low + High (H+L+H) values >= 95^th^ percentile compared with values <68^th^ percentile. The dashed lines indicate the point estimates for the whole study population.

**Figure 4.**
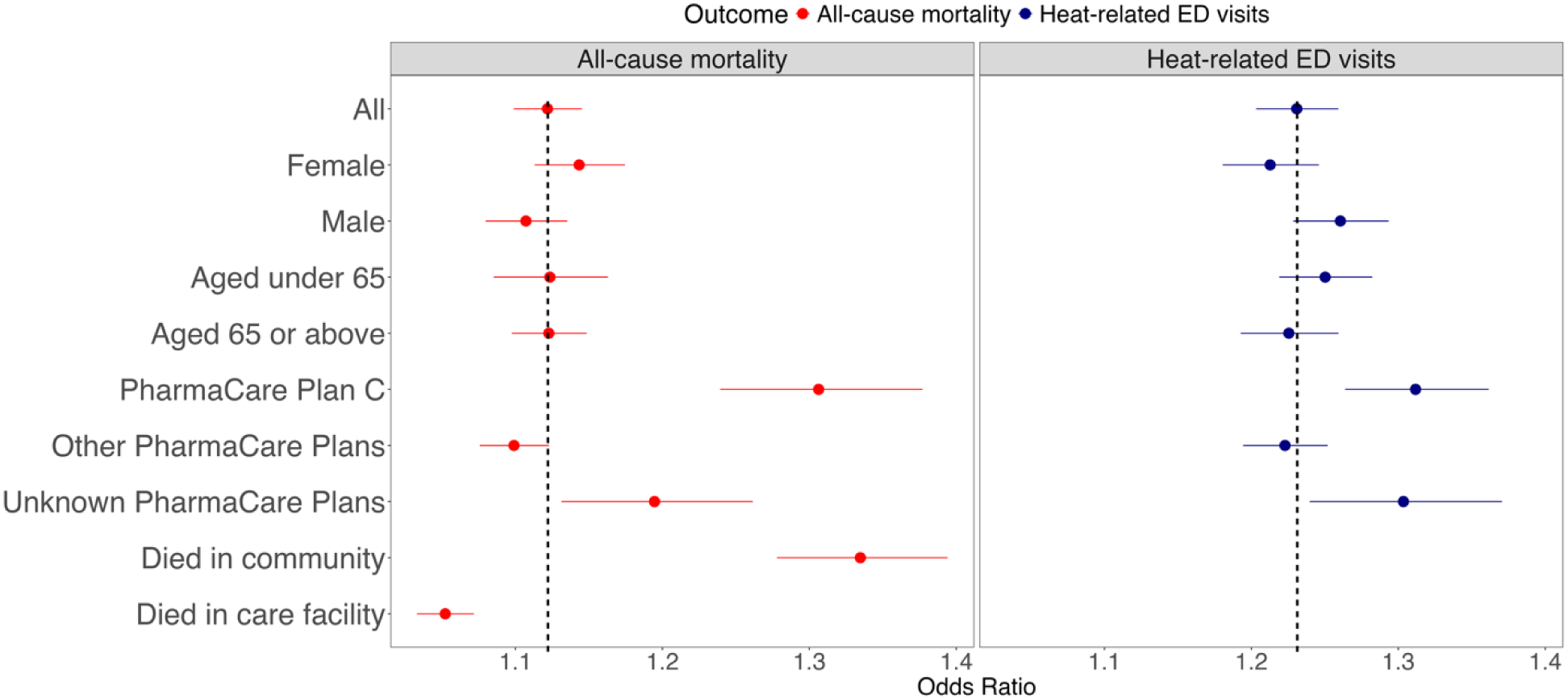
Odds ratios for all-cause mortality and heat-related emergency department (ED) visits, stratified by stratified by location of death, PharmaCare Plan status, sex, and age group, using White Rock temperature data with High + Low + High (H+L+H) values >= 95^th^ percentile compared with values <68^th^ percentile. The dashed lines indicate the point estimates for the whole study population.

Among the overall study population, the mortality OR was 1.12 [1.10, 1.15] and the heat-related ED visit OR was 1.23 [1.20, 1.26]. Several subgroups showed elevated susceptibility compared with the overall population. Individuals with schizophrenia had the highest mortality risk (OR: 1.37 [1.27, 1.48]) and the second-highest risk of ED visits (OR: 1.38 [1.31, 1.46]), whereas those with parkinsonism had the highest ED visit risk (OR: 1.47 [1.36, 1.59]) (Figure 3). In terms of demographic subgroups, individuals who died in the community had the highest mortality risk (OR: 1.33 [1.28, 1.39]) compared with those who died in care facilities (OR: 1.05[1.03, 1.07]).

Individuals enrolled in PharmaCare Plan C experienced elevated risks for both mortality (OR: 1.31 [1.24, 1.38]) and ED visits (OR: 1.34 [1.29, 1.40]). Those with unknown PharmaCare plan status also showed increased risks for mortality (OR: 1.19 [1.13, 1.26]) and ED visits (OR: 1.30 [1.24, 1.37]).

### Case study of current versus proposed approach

Using temperature data from White Rock, current heat alert criteria (29°C-16°C-29°C) were met during three distinct events between 2008-2024, including July 30-31, 2009, August 19-20, 2016, and June 26 to 30, 2021. The equivalent cumulative H+L+H threshold (≥74°C) identified these same events plus 40 additional dates (eFigure 2). Days meeting current criteria had an OR of 1.93 [1.83,2.05] for all-cause mortality and 2.17 [2.06,2.30] for heat-related ED visits. Days not meeting current criteria but meeting or exceeding the H+L+H value of ≥74°C had ORs of 1.08 [1.04, 1.12] and 1.21 [1.17, 1.26] for all deaths and ED visits, respectively. Across all days with H+L+H ≥74°C, the ORs were 1.35 [1.30,1.41] for mortality and 1.56 [1.50, 1.62] for ED visits (eFigure 5).

### Sensitivity analyses

Models with alternative specifications, including 8 degrees of freedom for temperature, a 7-day lag period, and 3 knots per year for long-term trends yielded results consistent with the reported findings (eFigure 3). Analyses using temperature forecasts issued three days in advance for Vancouver and Abbotsford showed similar minimum mortality and morbidity H+L+H values, and the exposure-response curves remained consistent for both absolute and relativistic H+L+H approaches (eFigure 4).

## Discussion

This study evaluated a proposed relativistic H+L+H metric for public heat alerts using population-level all-cause mortality and heat-related morbidity data for greater Vancouver, Canada. We found that relativistic H+L+H thresholds produced consistent exposure-response relationships across three weather stations in the region, while absolute temperature thresholds showed marked heterogeneity. The relativistic H+L+H approach also identified susceptible subpopulations, particularly individuals with schizophrenia, parkinsonism, those receiving income assistance and those who died in the community. The case study using current criteria and the equivalent H+L+H value demonstrated that the proposed approach captured additional days of elevated risk, suggesting the potential for improved population health protection.

The epidemiological rationale for cumulative heat exposure metrics reflects growing understanding of heat stress physiology and compound heat event risks under climate change^2,44,46–49^. Core body temperature regulation becomes progressively impaired when overnight cooling is insufficient to restore normal thermoregulatory function^50,51^. Recent studies in Europe and East Asia have demonstrated that high nighttime temperatures—often referred to as “tropical nights” when minimum temperatures exceed 20°C—are associated with increased mortality risk^49,50,52–54^. This physiological evidence supports the value of heat alerts that accommodate high overnight temperatures alongside somewhat lower daytime temperatures (e.g., 28°C-18°C-28°C versus 29°C-16°C-29°C in greater Vancouver).

Climate projections indicate that compound day-night heat events are becoming increasingly frequent and pose elevated mortality and morbidity risks due to limited overnight physiological recovery^2,46–49,55,56^. Research in China reported that compound heatwaves were associated with greater cardiopulmonary mortality risk than either daytime-only or nighttime-only events^46^. It found that by the 2090s, under moderate to high emission scenarios, mortality from compound heatwaves could rise 4.0–7.6 times compared with the 2010s, substantially exceeding projected increases from nighttime-only (0.7–1.9 times) and daytime-only (0.3–0.8 times) heatwaves^46^.

Similarly, research in East Asia found that days with hot nights were linked to 50% higher mortality risk compared to those without hot nights, with an estimated 3.68% (95% CI: 1.20– 6.17) of deaths attributable to hot nights by the end of 21^st^ century under a low-emission scenario^49^. The proposed H+L+H metric captures this cumulative burden by integrating both daytime heat intensity and overnight recovery potential, aligning with heat stress physiology and emerging epidemiological and climate evidence.

While global heat warning systems increasingly recognize multi-day heat exposure^57^, systems employing cumulative exposure metrics with relativistic thresholds remain limited. Most multi-day warning systems in European countries and North America use fixed absolute temperature thresholds to capture sustained heat exposure^22,24,25,28,58^, which may underestimate moderate-risk heat events, face challenges with spatial temperature variability, and require periodic recalibration as the climate changes. Some warning systems, such as the one in Spain^27^, use relativistic methods but typically apply them to fixed single-day temperature conditions.

Australia’s Excess Heat Factor (EHF) considers multi-day mean temperature exposure and relativistic thresholds but quantifies short-term heat anomalies using exclusively mean temperature deviations rather than deviations in cumulative daytime-nighttime exposure ^26,59^.

Our comparison with greater Vancouver’s current approach showed that the H+L+H metric identified 40 more heat risk days than the 29°C-16°C-29°C threshold. While days captured by the current system were associated with very risk, those missed by the current criteria but detected by the H+L+H approach still posed moderate risk. Although all heat alert systems must address public warning fatigue^57,60^, the cumulative and relativistic approach provides more nuanced information about risk, which can be integrated into risk-tiered systems already adopted by many countries ^24,26,28,33,57,58,61,62^.

Variability in absolute temperatures across greater Vancouver demonstrated that absolute H+L+H values produced different exposure-response functions within the same region, whereas the relativistic values produced consistent results. These spatial discrepancies highlight challenges in issuing heat alerts for climatically complex regions using absolute criteria. The ideal approach to heat alerts should use the most locally relevant exposure information to avoid biases in health effects estimation^63–67^. For example, two studies in the United States found that health effects of extreme heat vary within studied regions depending on the heat exposure method used and can be diluted due to exposure misclassification^66,67^.

To address the limitations of station-based exposure assessments, many researchers and meteorological service are moving towards fine-resolution gridded climate data^65,67–69^. However, even with spatially resolved estimates, assigning unique absolute heat alert thresholds to each grid cell remains operationally impractical. Therefore, our study addresses a fundamental challenge in heat warning systems for large and climatically diverse regions. Relativistic criteria can be applied across large geographies without periodic location-specific calibration, which is especially attractive for countries such as Canada given its land size and incoming transition toward high-resolution gridded forecasts and alerts. Moreover, relativistic heat warnings can use moving baseline data that account for climate change and local adaptation measures^70–73^. For example, heat warnings could be issued at H+L+H values over the local 95th percentile, based on the past 10 years of data.

The relativistic H+L+H approach showed higher temperature risks among some subpopulations, and generally higher risks for heat-related ED visits than for all-cause mortality. Risk was highest among those with schizophrenia, parkinsonism, substance use disorder, and COPD, as well as those receiving income assistance (Plan C) or lacking access to medical service (unknown plans) and those who died in the community. These findings align with existing literature that individuals with chronic conditions, low-income status, and limited access to cooling resources are particularly susceptible to extreme heat^21,74,75^. Most heat warning systems do not systematically incorporate susceptibility assessments for different population subgroups^22,57,76^. The ability of the relativistic H+L+H metric to detect different risks among subpopulations highlights its potential for informing personalized heat alerts or targeted public health interventions—shifting from broad, population-wide warnings toward more nuanced and equitable risk communication strategies^76^.

Overall, this study offers several strengths. First, it is situated in a heat-sensitive region—greater Vancouver—characterized by pronounced intra-urban temperature variability; by analyzing three stations spanning coastal to inland conditions, we explicitly address spatial heterogeneity that complicates heat alert operations in complex urban climates. Second, we leverage population-based outcomes from the provincial single-payer system, yielding comprehensive coverage of all-cause mortality and heat-related ED visits and enabling robust time-series analyses across a long observation window that includes multiple extreme heat events (notably the 2021 heat dome). Third, person-level linkages through PANDA to chronic disease registries and to a low-income proxy allow evaluation of susceptibility across clinically and socially vulnerable groups, directly informing equity-focused adaptations of alerting criteria. Finally, the work aligns with an active policy window: our partners from ECCC are modernizing heat alerts, and our H+L+H framework is already being tested in parallel with the current system, with full implementation anticipated by 2026.

A few limitations should be considered when interpreting these findings. First, the presence of heat warnings during many of the study dates may have affected exposure-response relationships through public health responses rather than representing pure temperature-health associations.

Second, we did not account for humidity or air quality, partially based on previous research in the region and the generally dry summer conditions that accompany hot weather in Vancouver^37^. Third, the pharmaceutical coverage proxy for low-income status may not capture all low-income individuals, due to the inherent limitations of administrative data sources. Finally, these findings may not be generalizable to regions with different climatic characteristics, population demographics, or healthcare systems. The relatively mild climate in greater Vancouver may limit the applicability of specific percentile thresholds to regions with more extreme summer conditions or different patterns of seasonal acclimatization. However, the methodological framework itself—applying relativistic approaches to cumulative exposure—should be broadly applicable, with region-specific calibration of threshold values.

## Conclusions

As climate change intensifies EHEs globally, robust and flexible heat health warning systems are critical for protecting public health across diverse geographic and climatic conditions. The relativistic H+L+H approach presented here lays an evidence-based framework for modernizing heat alerts in greater Vancouver and may be more broadly applicable in Canada and elsewhere. Operational testing in parallel with the current system was initiated in summer 2025 in partnership with the Meteorological Service of Canada, and full implementation is anticipated in the summer of 2026.

## Ethics statement

The study was approved by the University of British Columbia Research Ethics Board (Certificate number: H23-01938), and permission to access the health data was granted by the BCCDC PANDA team. Generative artificial intelligence (AI) was used solely for facilitating coding in the statistical analyses and was not involved in any other aspect of the work presented.

## Data Availability

Access to data provided by the Data Stewards is subject to approval but can be requested for
research projects through the Data Stewards or their designated service providers. The following
data sets were used in this study: Vital Statistics, NACRS, Chronic Disease Registries, PharmaNet. All inferences, opinions, and conclusions drawn in this publication are those of the author(s), and do not reflect the opinions or policies of the Data Steward(s). This data was provisioned under ISPs #21-051 and #24-066.

## Acknowledgments

We acknowledge the assistance of the Provincial Health Services Authority, BC Ministry of Health, and BCCDC staff involved in data access, procurement, and management. We gratefully acknowledge the residents of British Columbia whose data are integrated in the nb0107aa data mart. We also acknowledge ECCC for providing meteorological data. This work was funded by the British Columbia Ministry of Health.

## Data availability statement

Access to data provided by the Data Stewards is subject to approval but can be requested for research projects through the Data Stewards or their designated service providers. The following data sets were used in this study: Vital Statistics, NACRS, Chronic Disease Registries, PharmaNet. All inferences, opinions, and conclusions drawn in this publication are those of the author(s), and do not reflect the opinions or policies of the Data Steward(s). This data was provisioned under ISPs #21-051 and #24-066.

## Appendix

**eFigure 1.**
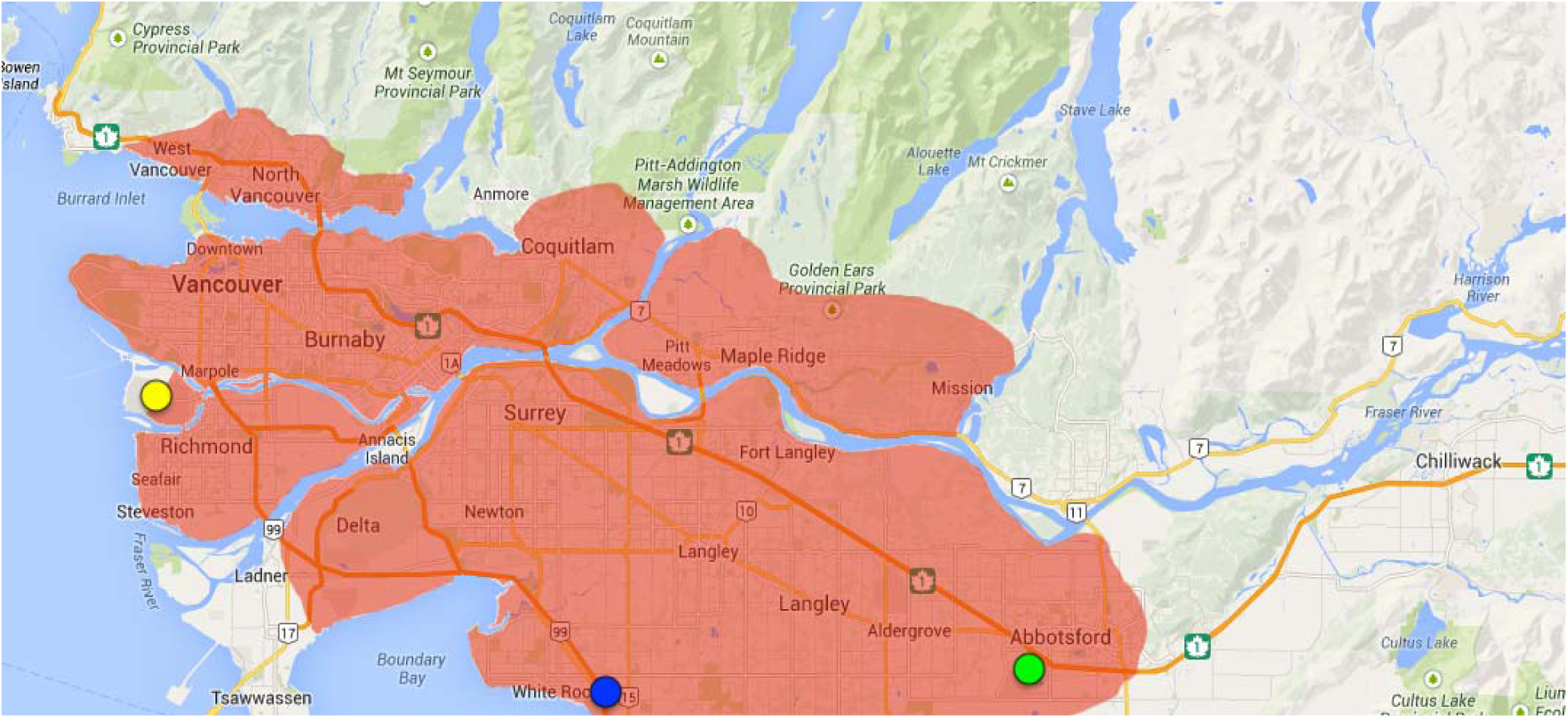
Map of the greater Vancouver region (in red) in British Columbia, Canada. Yellow marker is the Vancouver International Airport weather station, blue marker is the White Rock weather station, and the green marker is the Abbotsford International Airport weather station.

**eFigure 1.**
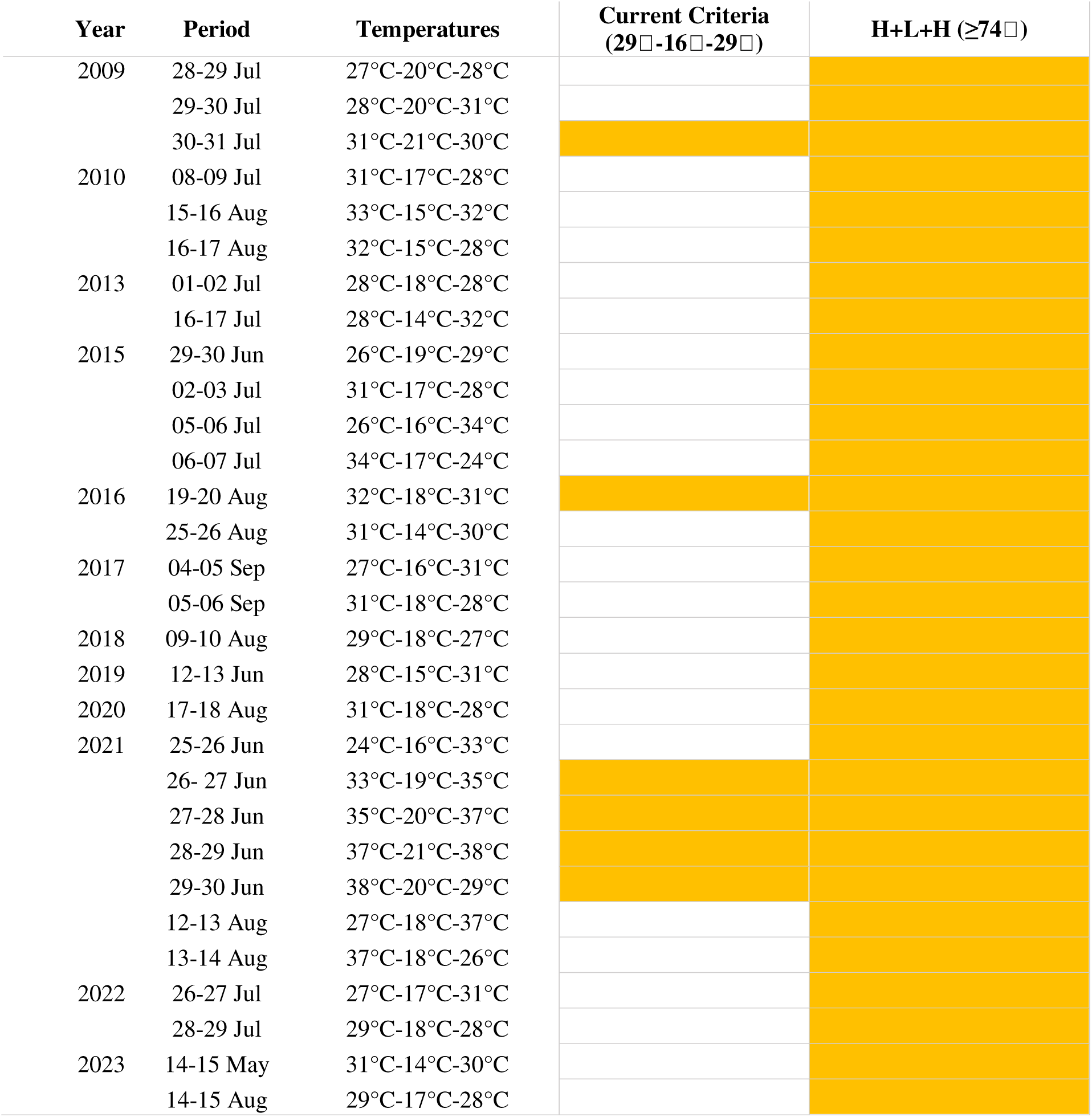
Comparison of dates meeting fixed versus cumulative heat thresholds in Metro Vancouver district between 2008 and 2024. Columns in orange are dates that meet or exceed the fixed or cumulative thresholds; Real Temperature Range has the actual temperature readings of High + Low + High (H+L+H).

**eFigure 2.**
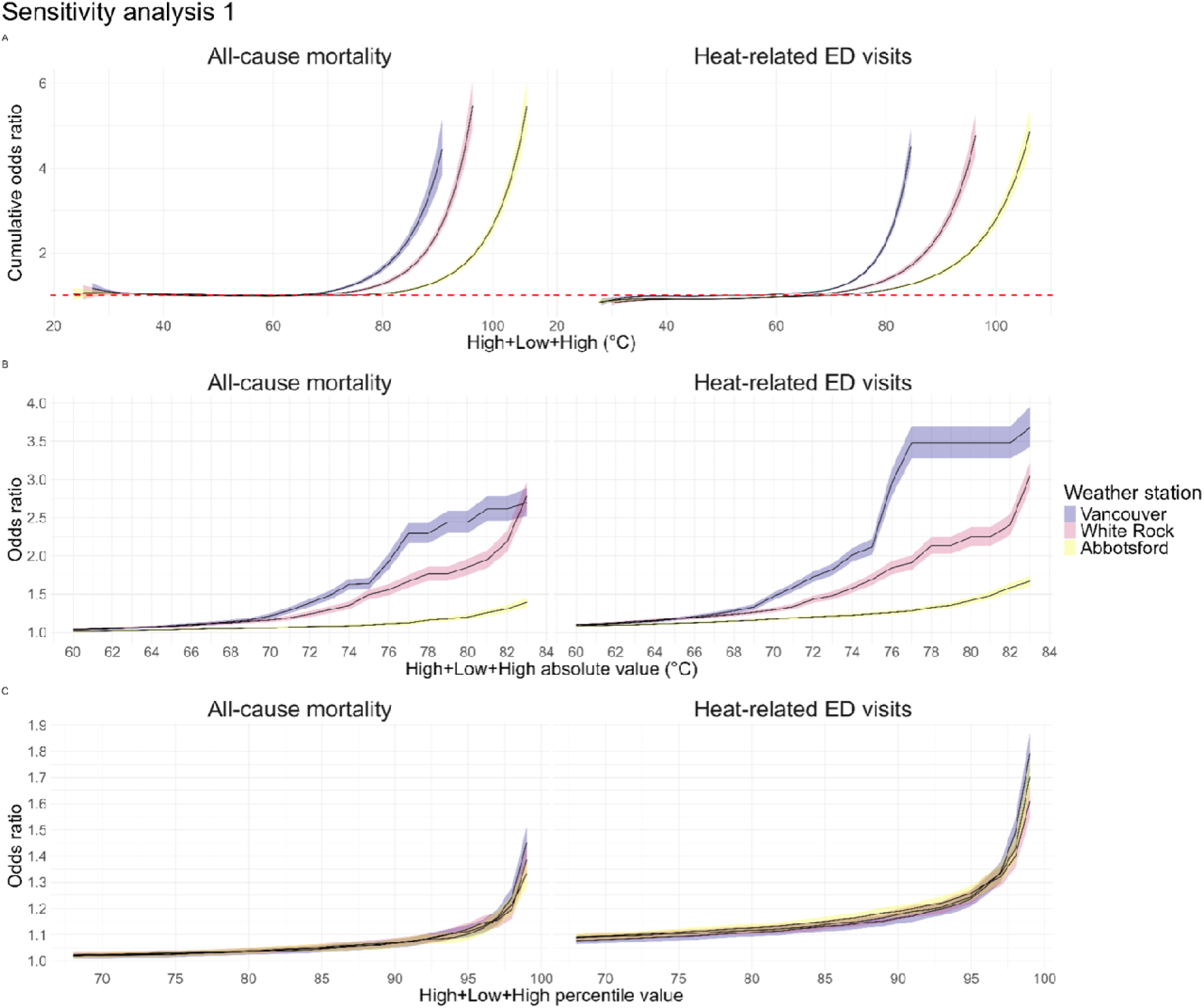
Sensitivity analysis 1: distributed lag non-linear models for identifying minimal mortality or morbidity temperatures used a 3-day lag, 8 degrees of freedom for temperature, and 3 knots per calendar year for long-term trends; negative binomial generalized additive models used 3 knots per calendar year for long-term trends for both absolute and relativistic HLH values.

**eFigure 3.**
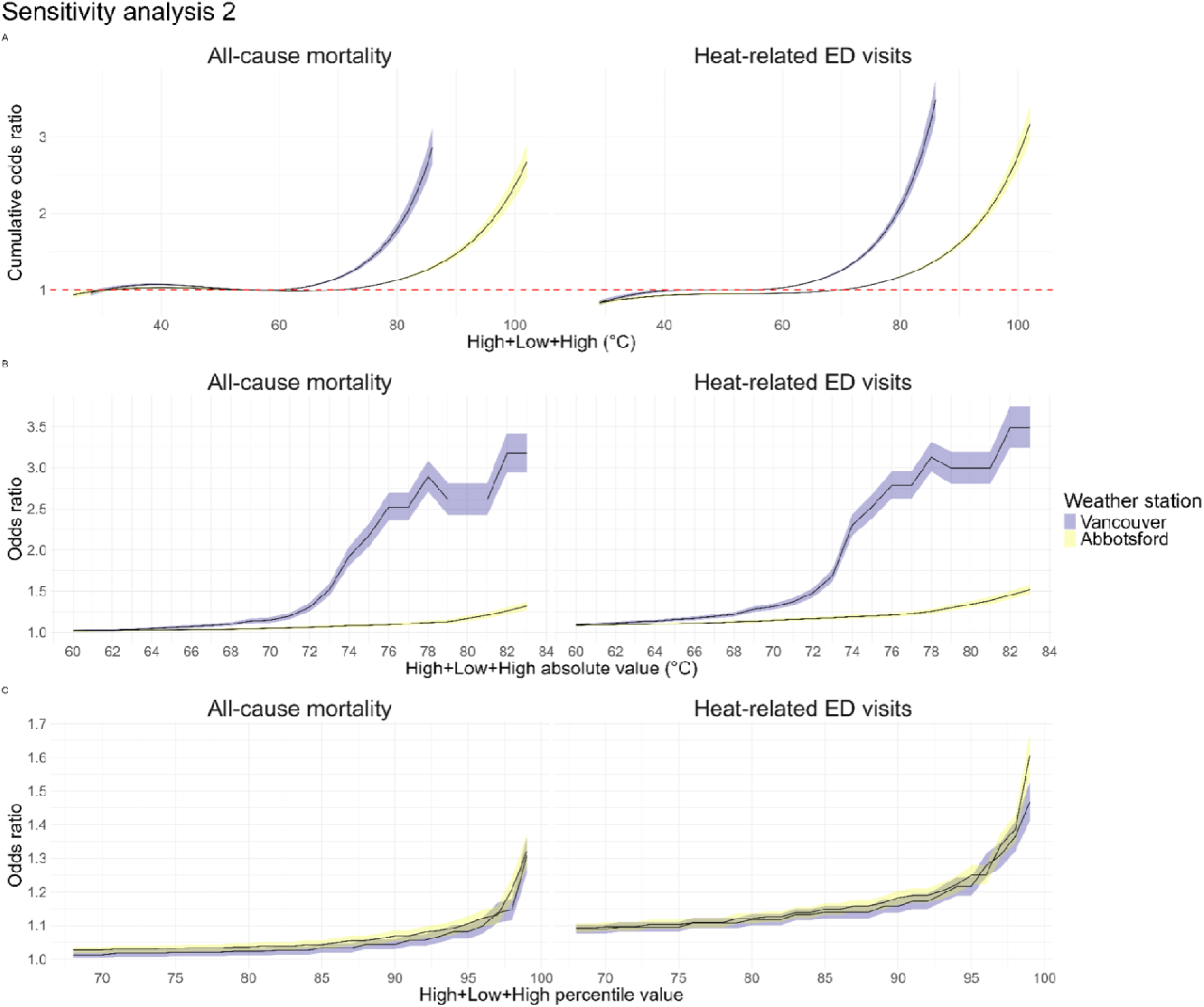
Sensitivity analysis 2: distributed lag non-linear models for identifying minimal mortality or morbidity temperatures and negative binomial generalized additive for both absolute and relativistic High + Low + High values, using temperature forecast data issued 3 days prior.

**eFigure 5.**
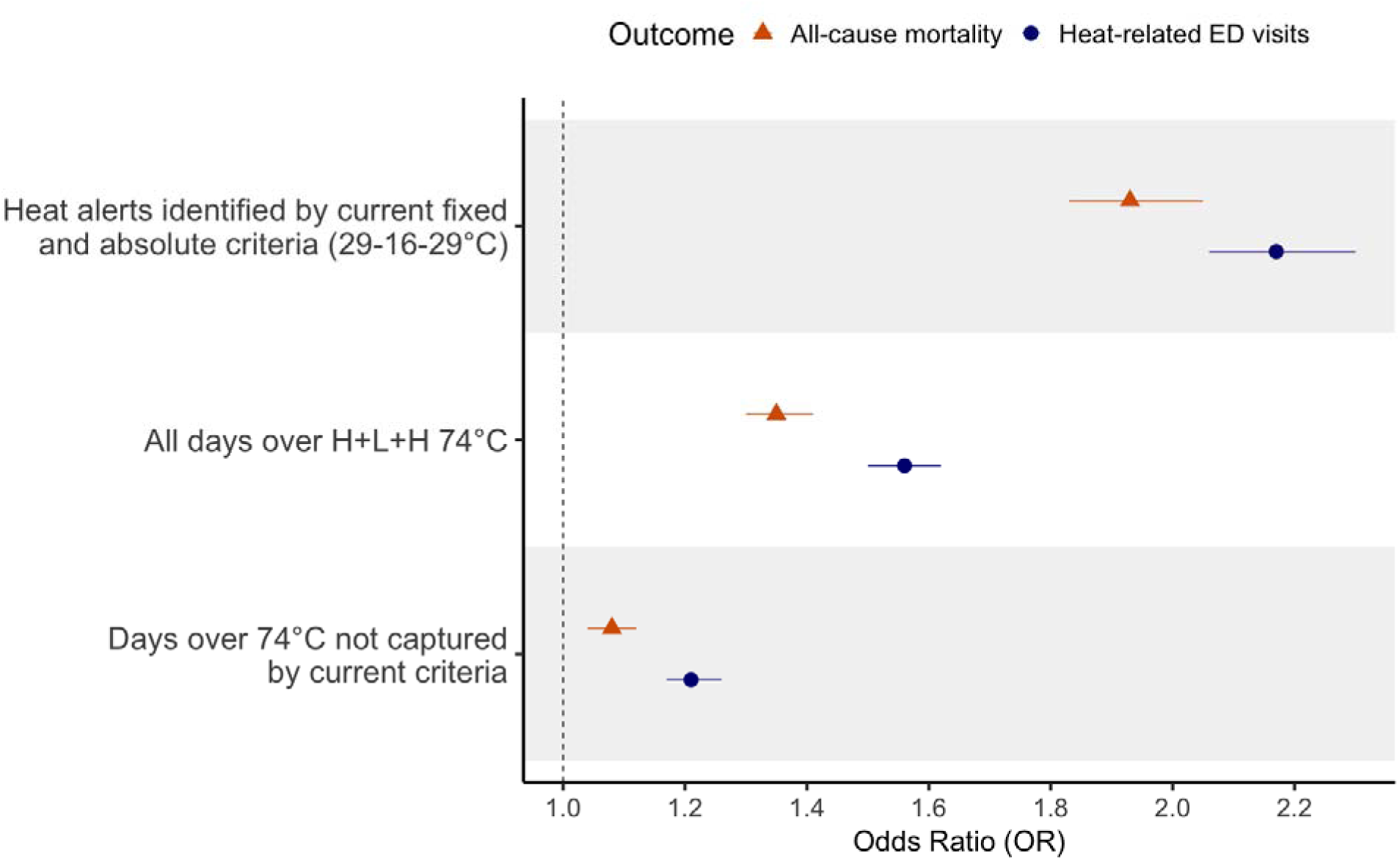
Odds ratios of all-cause mortality and heat-related ED visits on (1) days meeting current BC HARS heat alert criteria (29°C–16°C–29°C), (2) days missed by current criteria but captured by the cumulative H+L+H threshold (≥74°C), and (3) all days with H+L+H ≥74°C, relative to days below the minimum mortality baseline (H+L+H < 60°C). Estimates are from negative binomial generalized additive models with five knots per year, limited to the Metro Vancouver Regional District.

---

**eTable 1.** Mean (standard deviation) of High + Low + High (H+L+H), daily maximum, and overnight low temperatures in Vancouver, White Rock, and Abbotsford.

|  | **Vancouver** | **White Rock** | **Abbotsford** |
| --- | --- | --- | --- |
| Daily high | 20.4 (3.5) °C | 20.4 (3.6) °C | 22.4 (5.0) °C |
| Overnight low | 12.2 (2.8) °C | 12.2 (2.7) °C | 11.3 (2.9) °C |
| H+L+H | 52.9 (9.1) °C | 53.0 (8.8) °C | 56.1 (11.4) °C |

